# Clinical Characteristics of Coronavirus Disease 2019 (COVID-19): An Updated Systematic Review

**DOI:** 10.1101/2020.03.07.20032573

**Authors:** Zhangfu Fang, Fang Yi, Kang Wu, Kefang Lai, Xizhuo Sun, Nanshan Zhong, Zhigang Liu

## Abstract

**OBJECTIVE:** Clinical characteristics of novel coronavirus disease (COVID-19) have been described in numerous studies but yielded varying results. We aimed to conduct a systematic review on scientific literatures and to synthesize critical data on clinical traits of COVID-19 from its initial outbreak to pandemic.

**METHODS:** Systematic searches were conducted to identify retrospective observational study that contained clinical characteristics on COVID-19 through multiple databases. Two reviewers independently evaluated eligible publications. Data on clinical characteristics of COVID-19 were extracted and analyzed.

**RESULTS:** Seventy-two retrospective studies demonstrating the clinical characteristics of COVID-19 were included. A total of 3470 COVID-19 patients were synthesized to the final analysis in an unbiased manner. The most common symptom was fever (2878 [83.0%]), and 63.4% of the patients presented fever as onset symptom. There were 2528 [88.2%] of 2866 cases had abnormal lung findings on chest CT scan. Laboratory findings showed that 1498 [62.8%] of 2387 cases had lymphopenia, and 1354 [64.8%] of 2091 cases had an increased level of C-reactive protein (CRP). A total of 185 [11.5%] patients were admitted to intensive care unit (ICU) while the overall case fatality rate (CFR) was 3.7%. Compared to patients admitted outside of Hubei, China, those from Hubei had a significant higher ICU admission rate (21.9% vs. 2.5%, *p*<0.001). Also, CFR attributed to COVID-19 was significantly higher in Hubei than that of non-Hubei admissions (10.4% vs. 0.6%, *p*<0.001).

**INTERPRETATION:** This large patient-based systematic review presents a more precise profiling of the COVID-19 from its outbreak to current pandemic. Dynamic evolvements of COVID-19 are needed to be characterized in future studies.

## Introduction

In December 2019, a clustering pneumonia of unknown etiology occurred in Wuhan, Hubei Province, China^1^. The emerging disease has spread rapidly from Wuhan to other parts of China and to other countries^2^. Through a deep sequencing analysis, the virus responsible for this epidemic pneumonia has been designated as SARS-CoV-2 by International Committee on Taxonomy of Viruses (ICTV)^3^. The emerging disease caused by SARS-CoV-2 was named coronavirus disease 2019 (COVID-19) by WHO. By March 5, 2020, a total of 80,565 cases in China have been confirmed. Outside China, 14,768 cases of COVID-19 have been confirmed spanning in 85 countries, and the numbers are still on the rise^4^. What’s worse is that a total of 1,716 healthcare workers in China have been infected, among which 5 have died by February 11, 2020^5^. Thus, this infectious disease has become a serious global health concern.

Since the outbreak of COVID-19, a large number of articles have been published reporting epidemiologic and clinical characteristics of this emerging disease. Huang et al. first reported 41 cases of COVID-19 in which 66% patients had a history of exposure to Huanan Seafood Market, Wuhan. Common symptoms of the case series were fever (98%), cough (76%), and myalgia or fatigue (44%) with a case fatality rate (CFR) of 15%^6^. Chen et al. in a subsequent study found that 99 cases of COVID-19 patients admitted to the same hospital (Jin Yin-tan Hospital, Wuhan) had similar trends of symptoms and CFR (11%)^7^. Another case series analysis of 138 COVID-19 patients in Wuhan showed a relatively higher rate of fatigue symptom (69.6%) but lower CFR (4.3%) when compared to the above center^8^. Lastly, Xu et al.^9^ found in a multicenter-based retrospective study that clinical symptoms of patients in Zhejiang province are milder than those observed in Wuhan, China. Similarly, a nationwide multicenter study in China showed that patients with COVID-19 did not have a specific clinical symptom^10^. The clinical traits of COVID-19 are of great importance for the early identification of cases and for developing isolation and preventive strategies. However, there exist inconsistent findings of the clinical characteristics of COVID-19 among different studies, partly due to anthropogenic differences in patients enrolled and differences in sample size across different studies. Notably, variation in reporting descriptive data may lead to the misunderstanding of COVID-9 characteristics. In this updated systematic review, hence, we sought to address the heterogeneities among published retrospective studies and to synthesize the available data. We expect this critical review will provide insights to understand the clinical characteristics of COVID-19 in a more systematic manner.

## Methods

### Identification of Documents on COVID-9

Systematic searches were performed via the Medline database (PubMed) and Embase combining the terms (novel coronavirus OR 2019 novel coronavirus OR 2019-nCoV OR Coronavirus disease 2019 OR COVID-19 OR SARS-CoV-2). We also searched the database of Chinese Medical Journal full-text database (http://journal.yiigle.com/) for publications in Chinese using the above strategies. Searches were limited to publications from January 1, 2020 to March 1, 2020.

### Publication Selection

Two of the authors (Z. F. and F. Y.) independently screened searching results to determine inclusion or exclusion of the articles. Disagreements were modulated by consulting another author as adjudicator (Z. L.). We included retrospective observational studies as long as they contain clinical characteristics of COVID-19 associated illness. If the patients came from the same hospital with overlapping cases, we only selected the publication containing greatest number of cases. The following publication were excluded: reviews articles, meta-analysis, perspectives, comments, consensus documents, and publications that were in the neither English nor Chinese. Also, publications with highly suspected but not laboratory confirmed cases were excluded.

### Literature Quality Evaluation, Data Collection and Statistical Analysis

All included literatures were evaluated using the Newcastle-Ottawa Scale (NOS)^11^. The quality score of the literature ranged from lowest 0 to highest 9. Total quality score of 0-3, 4-6, and 7-9 indicated poor, fair, and good studies, respectively. Information on baseline demographic data, medical and exposure history, symptoms and signs, underlying comorbidities, laboratory findings, chest computed tomographic (CT) scans and CFR were recorded. Based on the diagnostic gold standard for COVID-19 (positive RT-PCR assay for SARS-CoV-2), we synthesized data on demographic and clinical parameters in an unbiased manner. The proportion of each parameter was calculated by the following formula: (actual patient counts)/ (total patients confirmed using the gold standard) × 100%. The χ2 test was used to compare the proportions. Statistical analyses were conducted using SPSS version 18 (IBM SPSS Statistics, IBM Corporation).

## Results

### General Information on the Systemic Review

A total of 1052 publications were retrieved using the search strategy. Seventy-two retrospective studies (including 22 case report, 20 case series, 3 case-control and 27 cross-sectional studies) that met the inclusion criteria were included in the final analysis **(Figure 1)**. Of the selected publications, 51 were written in English while 21 were written in Chinese. The general information on eligible publications can be found in **Table 1**. Ten publications reported COVID-19 cases from outside of China in USA^12^, Germany^13^, South Korea^14^, Vietnam^15 16^, Nepal^17^, Thailand^18^, Singapore^19^, Canada^20^ and Italy^21^. The remaining cases were from China covering 31 provinces or provincial-level municipalities. Additionally, nine reports specifically targeted children and the remaining focused mainly on adult patients (including 2 on pregnant women). Admission time for COVID-19 patients was from December 11, 2019 to February 14, 2020. Quality assessment of the literatures showed that cross-sectional studies achieved a good quality (median score, 7.0) while case report/series were fair (median score, 6.0).

**Table 1.**
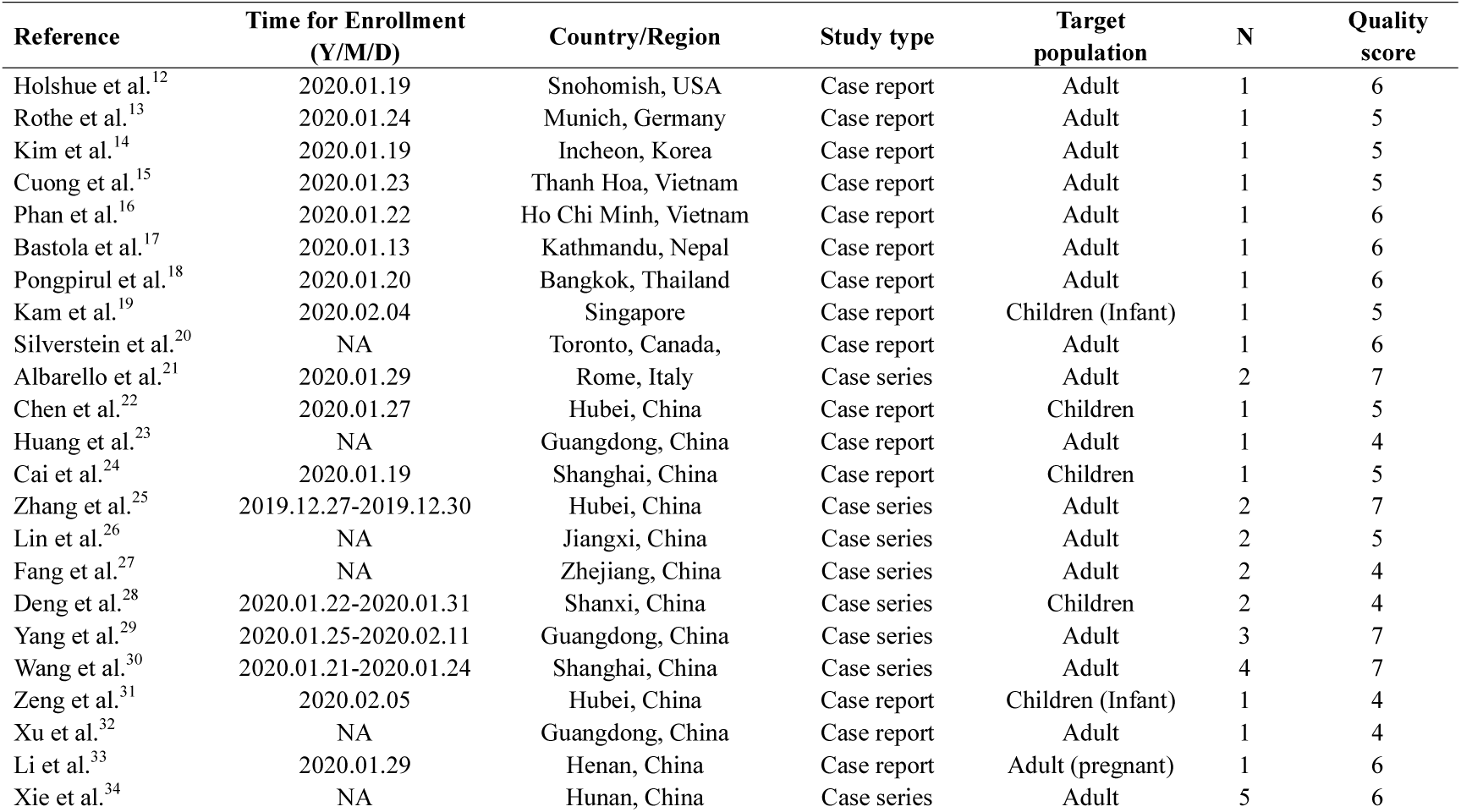
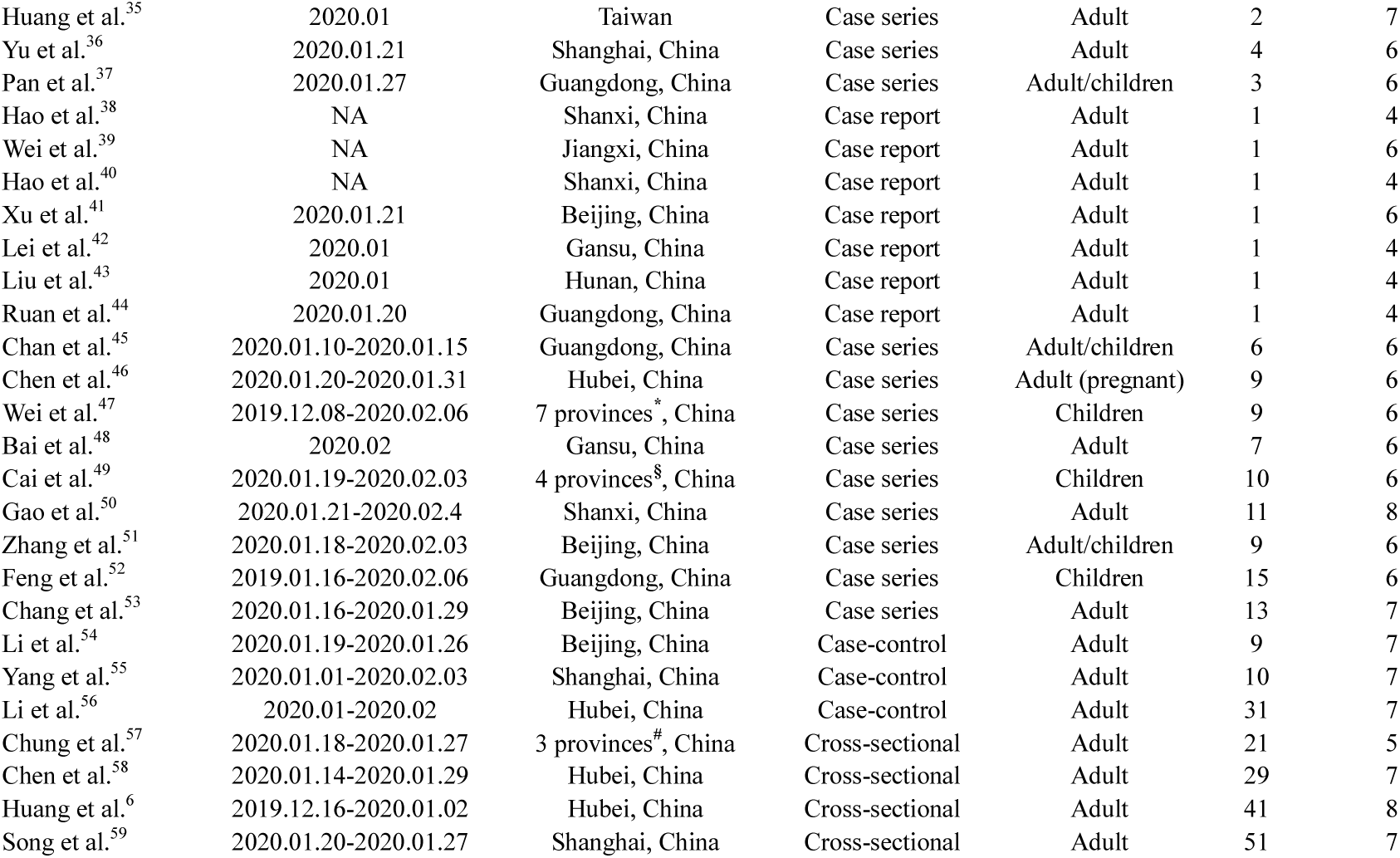
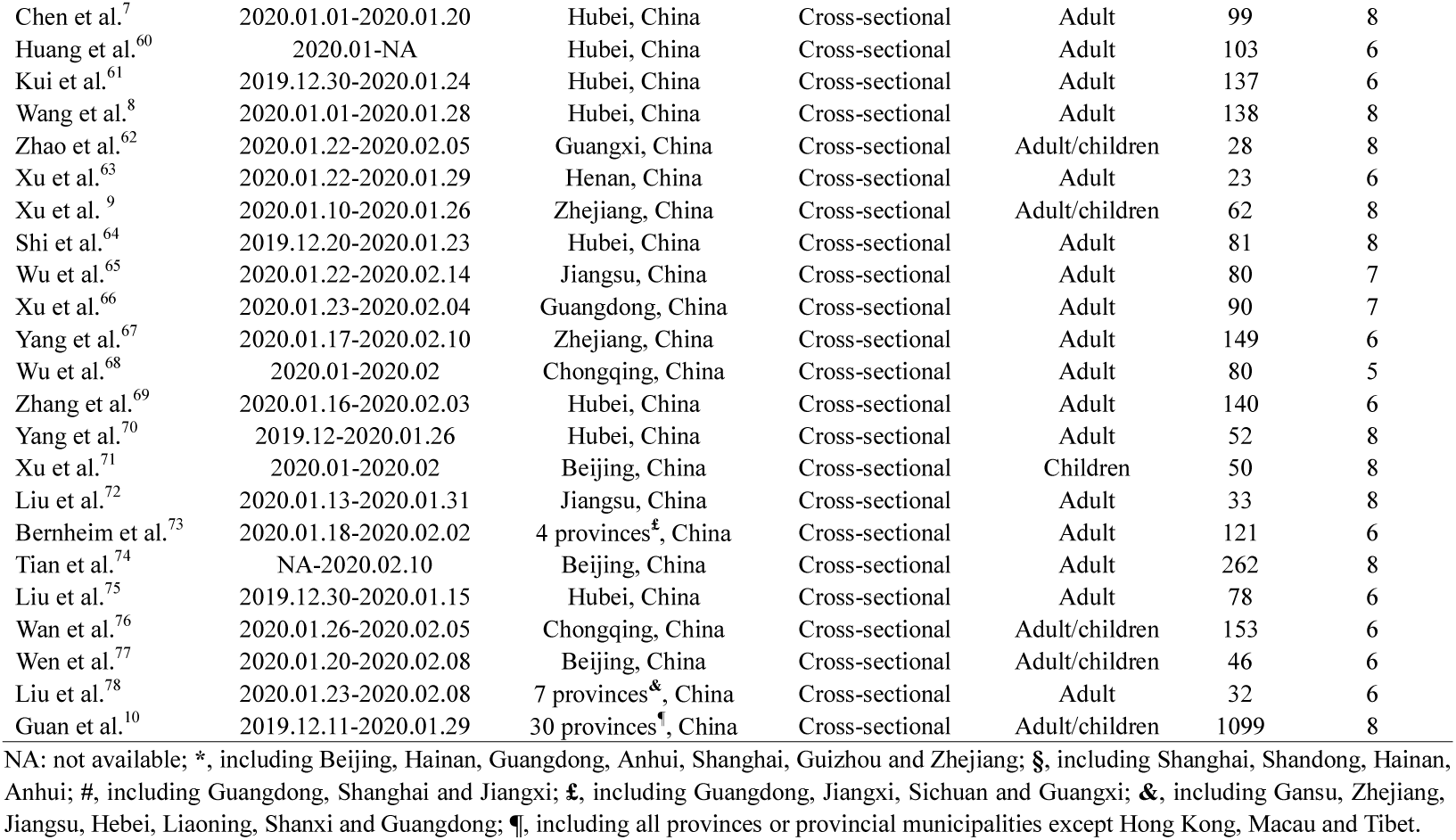
General information of the identified literatures.

**Figure 1.**
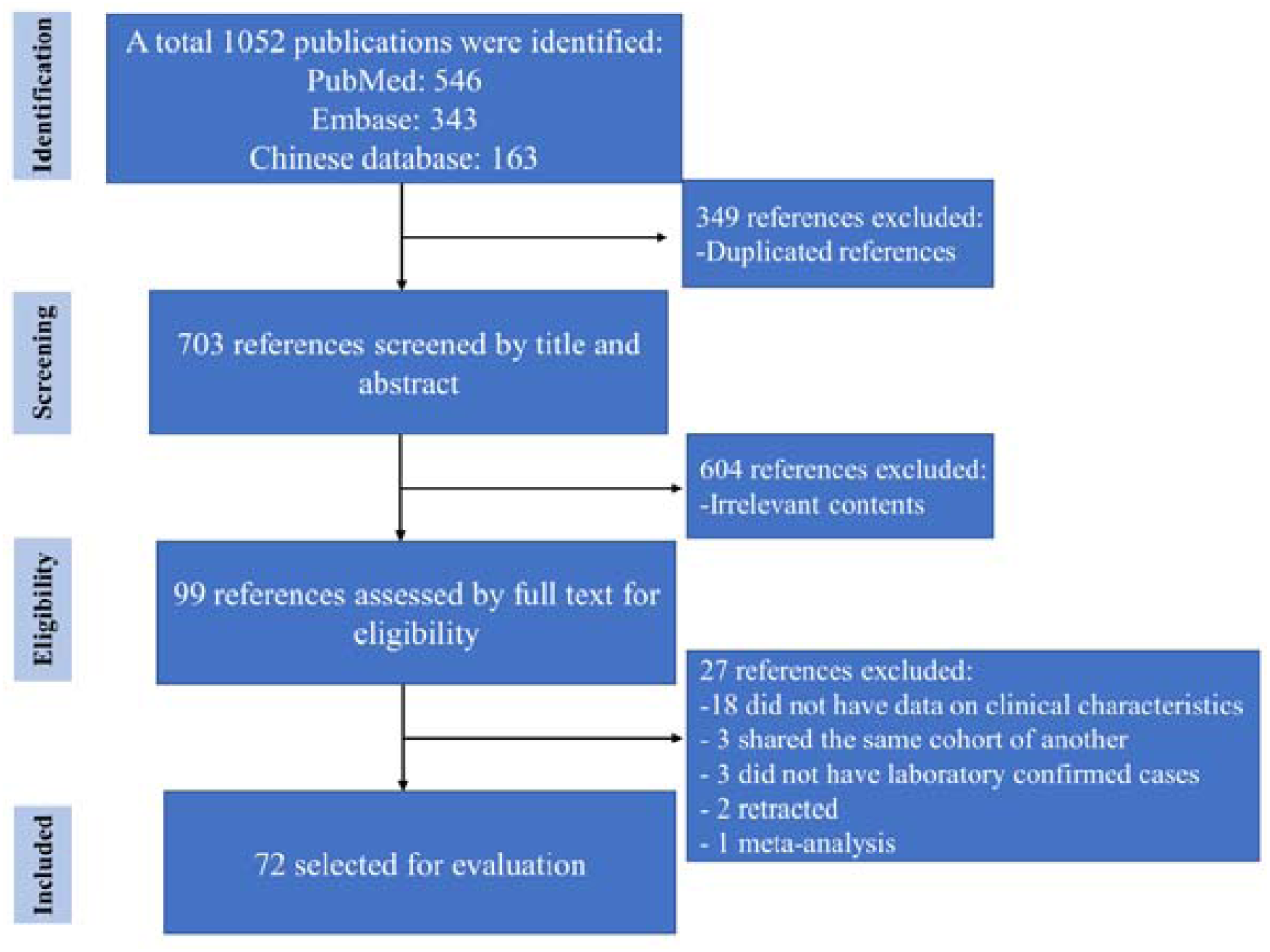
PRISMA flow chart of literature searching and selection.

### Demographics and epidemiology of Selected Patients

A total of 3,470 COVID-19 patients were included, of which 3,468 confirmed cases were based on the positive SARS-CoV-2 on RT-PCR assay while two cases^25^ were diagnosed according to coronavirus antibody detections. The age of the COVID-19 patients ranged from 17 days to 92 years old. Male gender accounted for 1822 (52.6%) of the selected patients. Among 1,573 patients who were asked for smoking status, 208 (13.2%) were self-reported smokers. Through a synthesized analysis, a total of 2182 cases (76.0%) had a history of transmission exposures, *i*.*e*., the patients were either Wuhan residents or travelled to Wuhan within the past 14 days.

### Clinical Characteristics of COVID-19

The most common symptom of COVID-19 was fever (2878 [83.0%]), followed by cough (2102 [61.0%]), fatigue (942 [37.9%]), sputum production (720 [28.7%]), dyspnea (412 [14.5%]), muscle aches (477 [18.6%]). Less common symptoms were headache (318 [11.8]), sore throat (289 [14.0%]), gastrointestinal symptoms (anorexia, nausea or vomiting account for 185 [8.9%]) and upper airway symptoms (rhinorrhea, sneeze or nasal congestion account for 162 [7.6%]) and diarrhea (165 [6.1%]) (**Figure 2**). Further analysis showed that only 1522 [63.4%] of the patients had fever as their onset symptom.

**Figure 2:**
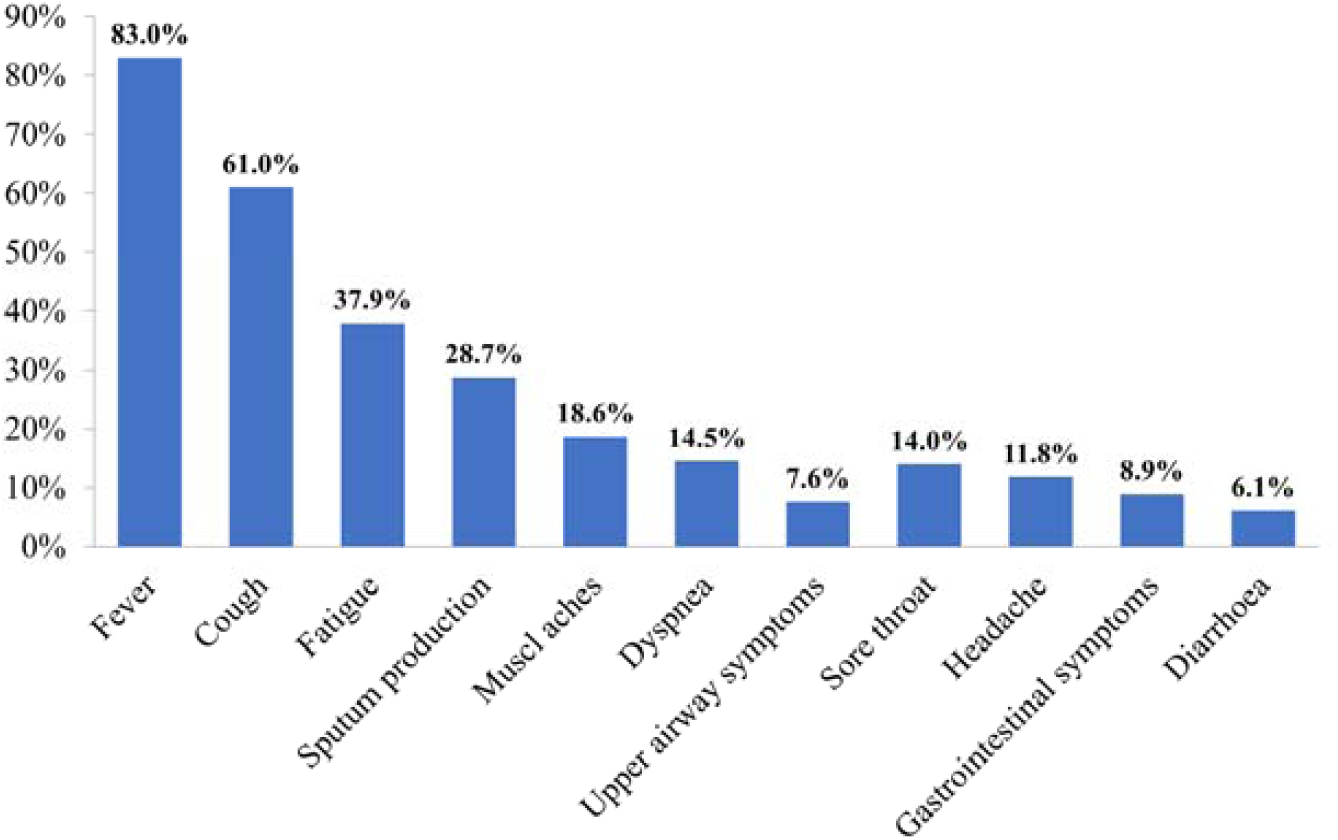
Clinical symptoms of patients with COVID-19.

Of the selected patients, 888 [31.5%] cases co-existed with underlying chronic disorders, such as hypertension [13.3%], diabetes [7.3%], chronic obstructive pulmonary disease (COPD) [1.4%], cardiovascular and cerebrovascular diseases [8.3%], malignancy [1.5%], chronic liver disease [2.1%] and chronic kidney disease [0.7%] (See Table 3 for details). Common complications of COVID-19 included acute respiratory distress syndrome (ARDS) (136 [8.9%]), shock (29 [2.2%]) and acute renal failure (30 [2.1%]).

Radiological findings revealed that 2528 [88.2%] of 2866 cases had abnormal presentations on chest CT **(Figure 3A)**, with either bilateral or unilateral ground-glass/consolidative pulmonary opacities. Laboratory blood tests showed that 1498 [62.8%] of 2387 cases had lymphopenia (lymphocyte count < 1.0 × 10^9^/L), while 1354 [64.8%] of 2091 cases presented an increased level of C-reactive protein (CRP) **(Figure 3 B-C)**.

**Figure 3:**
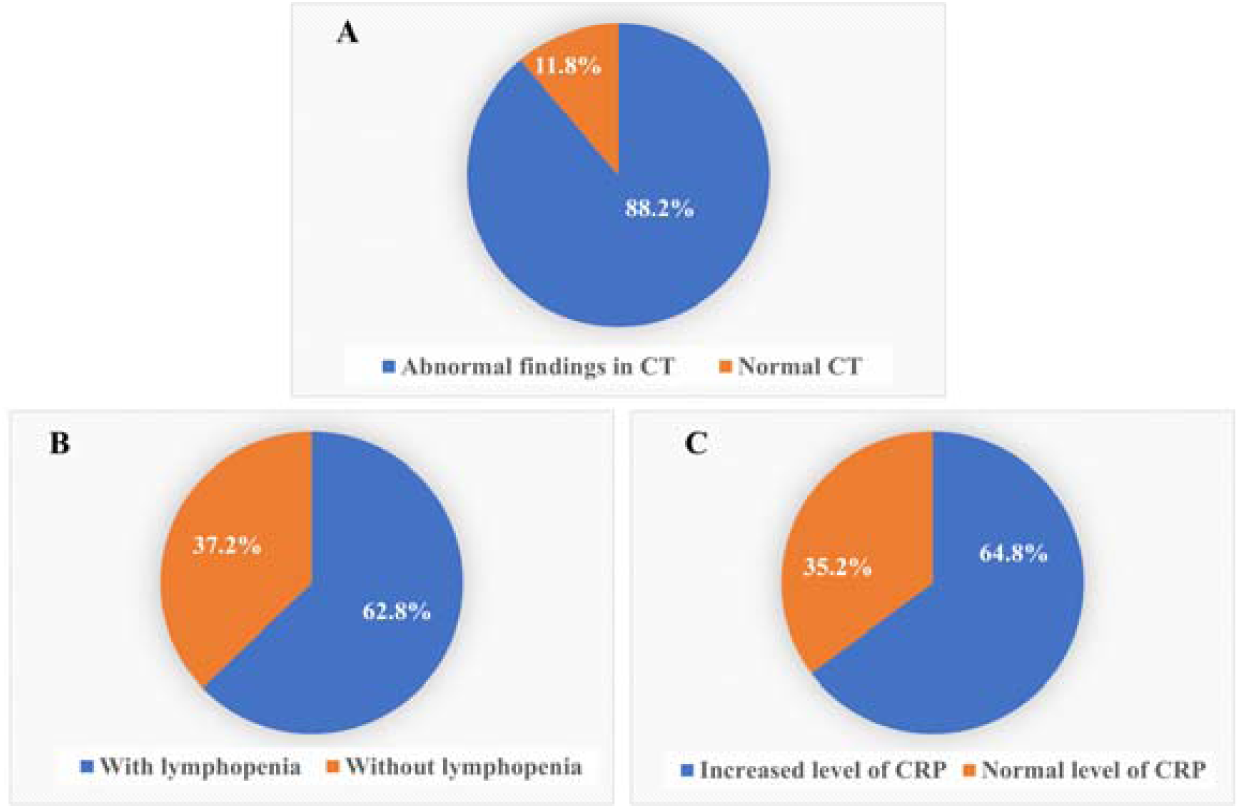
Findings of radiography (A) and laboratory blood tests (B, C) in patients with COVID-19.

### Other Outcomes

Collectively, a total of 185 [11.5%] patients were admitted to intensive care unit (ICU) while the overall CFR was 3.7% (**Table 2)**. Our further analysis showed that COVID-19 patients admitted in Hubei province (Wuhan in particular) suffered from a significant higher ICU admission rate than that outside of Hubei, China (21.9% vs. 2.5%, *p*<0.001). Also, CFR attributed to COVID-19 in Hubei province, China was significantly higher than that in non-Hubei, China (10.4% vs. 0.6%, *p*<0.001).

**Table 2:** Summary of the demographics and clinical characteristics of patients with COVID-19.

| **Demographics** |  |
| --- | --- |
| Total number of cases | 3470 |
| Male% | 1822/3467, 52.6% |
| Age (range) | 17-day to 92-year |
| Smokers% <sup>#</sup> | 208/1573, 13.2% |
| Transmission exposures% <sup>*</sup> | 2182/2873, 76.0% |
| **Coexisting underlying disorders (888/2818, 31.5%)** |  |
| Diabetes | 206/2818, 7.3% |
| Hypertension | 376/2818, 13.3% |
| Cardiovascular and cerebrovascular diseases | 233/2818, 8.3% |
| Chronic obstructive pulmonary disease (COPD) | 40/2818, 1.4% |
| Chronic liver diseases | 58/2818, 2.0% |
| Chronic renal diseases | 20/2818, 0.71% |
| Malignancy | 43/2818, 1.5% |
| Digestive system disease | 32/2818, 1.1% |
| Other chronic diseases<sup>§</sup> | 148/2818, 5.3% |
| **Complications** |  |
| ARDS | 136/1526, (8.9%) |
| Shock | 29/1347, (2.2%) |
| Acute renal failure | 30/1399, (2.1%) |
| **ICU admission** | 185/1612, (11.5%) |
| **Case fatality** | 100/2682, (3.7%) |

Notes: #, current smokers and ex-smokers; \*, being Wuhan residents or ever travelled to Wuhan within 14 days; §, Including thyroid disorders, HIV, endocrine system disease, nervous system disease, etc.

## Discussion

This review analyzed clinical data on COVID-19 patients enrolled retrospectively in published studies. These patients were admitted to hospitals between December 11, 2019 and February 14, 2020, which covered a period from COVID-19 outbreak to its global pandemic. To our knowledge, this updated systematic review analyzed the largest number of cases demonstrating the clinical characteristics of COVID-19, which covers ten countries. As of March 5, 2020, there have been 95,333 confirmed cases distributing over 86 countries^4^. The rapid and wide spreading of the infection has clearly shown the pandemic potential of COVID-19, to which we should pay great attention.

Through literature searching and data extraction, we included 3,470 confirmed cases of COVID-19 based on the positive assays of SARS-CoV-2 on RT-PCR or antibody detection. The present study showed that the infected patients aged from 17-day to 92-year old. A recent report by Chinese Center for Disease Control and Prevention (China CDC) have shown that 86.6% the patients aged 30-79 years^5^. This study together with China CDC report indicated that the general population, regardless of age, is susceptible to SARS-CoV-2 infection. The general ratio of male to female gender of the present study (1.10:1) is also similar to China CDC report (1.06:1), indicating that COVID-19 may not have a gender predisposition. Although 13.2% of the selected patients had a history of smoking, we could not draw the conclusion that smokers are less susceptible to the viral infection than non-smokers, as the patients included in our analysis were from a sample pool that was not representative the general population by smoking status.

This systematic review showed that most of the patients (76.0%) had a history of Wuhan-related exposures. This is of great importance for early quarantine for the subjects with emerging infectious disease when specific treatments are not available. Actually, we have learned from the SARS outbreak 17 years ago that early identification, early isolation and early management would lead to the stop of viral transmissions from human to human^79^. Collectively, there were 36% of the COVID-19 patients absent from fever as the onset symptom. In this case, such patients may have been ignored at the early stage if we focused heavily on fever examination for initial screening. Our composite analysis showed that fever remains the most common symptom (83.0%) in patients with COVID-19. However, the proportion of fever is somewhat lower than that of other coronavirus related respiratory illness, such as SARS (100%)^80 81^ or middle east respiratory syndrome (MERS) (98%)^82^. Similarly, the accompanied symptoms of dyspnea (14.5%) and diarrhea (6.1%) are relatively less common in patients with COVID-19 than those seen in SARS and MERS^87^. More importantly, this study revealed an overall CFR of 3.7%, which was quite similar to that reported by the WHO official statistics as of March 5, 2020 (CFR 3.7%, 3,015 died of 80,565 cases)^4^. Nevertheless, the CFR of COVID-19 was much lower than that of SARS (9.6%)^83^ and MERS (37.1%)^84^. By far, the mechanisms underlying the varying symptoms and CFR for these three coronavirus-infected diseases are not fully understood. One reason may be that there were still some COVID-19 patients being treated in hospitals at the time of the manuscripts submitted, so the outcome (death or recovery) is not known yet. Additionally, we suppose that the varied tropism as well as virulence of these three coronaviruses may in part account for the discrepancies, which warrants further investigations.

According to the seven version Guidelines for the Diagnosis and Treatment of Novel Coronavirus Infection^85^, typical CT findings of early COVID-19 are characterized by multiple small patchy shadows and then developed into bilateral pulmonary ground-glass or consolidative opacities. In our summary, most of the patients (88.2%) had abnormal findings in the lungs with chest CT scan. In some early cases of COVID-19, clear evidence of pneumonia has been shown on HRCT while X-ray radiographic findings are still normal^51^. Thus, chest CT scan should be routinely recommended for the early identification of COVID-19. Routine blood tests showed that 62.8% of COVID-19 patients had lymphopenia, of which the level is relatively lower than that of SARS^86^. Similarly, a large proportion (64.8%) of COVID-19 patients had an increase level of CRP. Taken together, a combination of the epidemiologic history with auxiliary examinations, *e*.*g*., chest CT scan and blood tests, will improve the diagnostic accuracy for COVID-19.

Our further analysis revealed that patients admitted in Hubei province (Wuhan in particular) are facing much more severe situations (higher ICU admission rate and CFR) than those diagnosed in other parts of China. Reasons for the tough situations being caught in Hubei province might be as follows. Firstly, the human-to-human transmission of COVID-19 had not been cutting down at its early stage in Wuhan, Hubei province. Second, when encountering such a large number of infected patients, a limited healthcare workforce in Wuhan could not support to treat all patients at their mild or moderate stage. This might result in a higher rate of ICU admission or CFR. Since substantial number of patients as well as healthy residents have been kept in lockdown of Wuhan, great efforts are urgently needed to cutdown the virus transmission from COVID-19 patients to local healthy residents.

This systematic review has some limitations. First, due to the retrospective nature of selected literatures, not all clinical characteristics have been well documented. This have resulted in an inconsistence of the total numbers of each item being calculated. Second, some of the selected patients may not have discharged before the studies were finalized. This might have affected the clinical outcomes of COVID-19 to some extents. Third, since the disease is still ongoing, newly published data on COVID-19 might not be included at the time of manuscript submission. Nevertheless, we have summarized the largest number of patients demonstrating the clinical characteristics of COVID-19 in the present study. Finally, we could not determine the incubation periods of COVID-19 due to heterogeneity across studies in reporting the timeline of cases.

In summary, COVID-19 represents an emerging acute respiratory infection with various clinical presentations that share similarities as well as discrepancies with SARS and MERS. Combining clinical presentations with radiographic findings as well as blood tests may add value in early diagnosis and managements. Dynamic changes of clinical features in the course of COVID-19 are needed to be characterized in future studies. Since specific treatments are not available at the moment, urgent efforts should be taken to explore for this emerging disease.

## Data Availability

Data extraction files are available on request from the corresponding author (ZGL). The paper’s guarantor (ZGL) affirms that the manuscript is an honest, accurate, and transparent account of the study being reported; that no important aspects of the study have been omitted.

## Acknowledgement

We would like to thank Prof. Junfeng (Jim) Zhang from Duke University and Dr. Mei Jiang from Guangzhou Institute of Respiratory Health for their kind help in refining the manuscript.

## Contributors

ZF and FY contributed equally to this work. ZF, FY, and KW undertook the review. All authors contributed to the conception of the work and interpretation of the findings. ZF, FY, and ZGL drafted the manuscript. All authors read the manuscript and approved the final version. NSZ and ZGL acts as guarantor. The corresponding author attests that all listed authors meet authorship criteria and those did not meet the criteria have been omitted.

## Ethical approval

Not required.

